# The impact of the first wave of COVID-19 on those with lifelong conditions: a case study of congenital heart disease

**DOI:** 10.1101/2021.01.13.21249447

**Authors:** Jo Wray, Christina Pagel, Adrian H. Chester, Fiona Kennedy, Sonya Crowe

## Abstract

**Objectives:** Globally, health care systems have been stretched to the limit by the COVID-19 pandemic. Significant changes have had to be made to the way in which non-COVID-19 related care has been delivered. Our objective was to understand, from the perspective of patients with a chronic, life-long condition (congenital heart disease, CHD) and their parents/carers, the impact of COVID-19 on the delivery of care, how changes were communicated and whether health care providers should do anything differently in a subsequent wave of COVID-19 infections.

**Design and setting:** A series of asynchronous discussion forums set up and moderated by three patient charities via their Facebook pages.

**Participants:** Patients with CHD and parents/carers of patients with CHD.

**Main outcome measures:** Qualitative responses to questions posted on the discussion forums.

**Results:** The forums ran over a 6-week period and involved 111 participants. Following thematic analysis of the transcripts, we identified three themes and ten subthemes related to individual condition-related factors, patient-related factors and health professional/centre factors that may have influenced how patients and parents/carers experienced changes to service delivery as a result of COVID-19.

**Conclusions:** Our findings, whilst collected in relation to patients with CHD, are not necessarily specific to this population and we believe reflect the experiences of many thousands of people with life-long conditions in the UK. Drawing on what participants told us in the discussion forums, we have developed recommendations related to communication, service delivery and support during the pandemic that would, we think, improve patients’ experience of care and, potentially, their outcomes. Although the data were collected specifically in relation to COVID-19, a number of these recommendations are relevant to the wider delivery of care to patients with chronic underlying health conditions and reflect principles of good communication and service delivery.

## Background

Since late 2019, COVID-19 has spread rapidly around the world, reaching official pandemic status in March 2020.(1) The speed with which the virus has spread and the trail of physical and psychological illness, death and economic hardship have been extensively documented in the medical and everyday press. Vast amounts of resources have been ploughed into researching the transmission, disease trajectory and risk factors associated with COVID-19. Adults with underlying health conditions have been identified as being at increased risk of developing severe and fatal disease, particularly those with pre-existing hypertension and coronary heart disease.(2) In contrast to the adult population, severe COVID-19 infection in children is rare but there is a lack of comprehensive data on how children with underlying health conditions are affected by COVID-19.(3)

Globally, health care systems have been stretched to the limit and significant changes to the way in which non-COVID-19 related care has been delivered have had to be implemented. The periods of lockdown imposed in many countries and the cessation of non-essential face to face patient contact have necessitated rapid adjustments and adaptation to new ways of delivering and receiving care. Concerns have been raised about the impact of these changes in terms of delayed diagnosis of other health conditions,(4) delays in seeking treatment,(5) cancellations of treatment,(6) greater non-adherence to medical therapy (7) as well as increased mental health problems.(8) Whilst health professionals and the media have been vocal about these potential consequences, far less has been heard from the patients and their families who are being directly affected.

Congenital heart disease (CHD) is one example of a chronic, life-long condition with a spectrum of severity from mild to life-threatening. Both paediatric and adult patients typically require regular follow-up with specialist CHD professionals and tests of cardiac function are a cornerstone of follow-up. But, as with other patient groups, services for patients with CHD have seen significant and abrupt changes over the last 9 months. In common with many other underlying health conditions, it is currently unclear what risk COVID-19 presents to a patient with CHD. As part of a larger study commissioned by the NHS to develop new ways of measuring the quality of CHD services for both children and adults,(9) we set out to understand, from the perspective of patients and parents/carers, the impact of COVID-19 on the delivery of care, how changes were communicated and whether health care providers should do anything differently in a subsequent wave of COVID-19 infections. Our belief was that the learning and recommendations arising from this work would also be generalisable to the larger population of children and adults receiving care for other chronic health conditions.

## Methods

### Design

A qualitative approach underpinned by an interpretivist framework was used, in which online discussion forums were employed to elicit participant (patient or parent/carer) views.

### Patient and public involvement (PPI)

A patient co-researcher (AC) was involved with each stage of the project, including data analysis and revising drafts of the manuscript. AC also led a PPI group set up as part of the larger overarching study (comprising three adults with CHD and one grandparent of a child with CHD), who reviewed the forum questions and the findings prior to submission. The online discussion forums were moderated by three patient organisations, each of which contributed to the content and format of the questions. A summary of the results will be disseminated to all three charities for publication on their website and will also be disseminated to CHD services nationally via the Adult CHD specialist nurse network and NHS England.

### Participants and data collection

The Children’s Heart Federation, Little Hearts Matter and the Somerville Foundation, all of which are national UK charities dedicated to the support of patients with CHD and their families, facilitated and moderated one or more closed, anonymous, asynchronous online discussion groups via their Facebook pages, following an approach that we have successfully used in previous work.(10, 11) We specifically chose these three charities because we wanted to collect views across age ranges (parents of younger children, teenagers and adult patients with CHD) and from those with complex and less complex CHD. Questions were developed by the authors and the content and language revised based on feedback from the charity representatives and PPI group. The charities recommended that separate forums should be facilitated for adult patients with CHD, teenage patients with CHD and parents/carers of children and young people with CHD. Each charity advertised the discussion forums on their home web page and potential participants were directed to the charity’s Facebook page where they were able to access further information about the purpose of the forum, how it would be facilitated and the governance surrounding it. People interested in participating were asked to provide some basic demographic information (age, gender, ethnicity, name of CHD defect, location of home and specialist service, relationship to the person with CHD, and age of person with CHD (for parents/carers)). Having completed this information, they were directed to the appropriate closed Facebook group, depending on participant group, where they were able to respond to the posted questions. The research team provided each charity with the agreed questions at the start of the process and the charity determined when new questions should be posted or any prompts introduced, based on responses. The forums took place over a 6-week period, from August 2020 to September 2020. Questions were very similar for each participant group and each charity, with small revisions to wording to reflect the respondent group (e.g. patient- or carer-relevant wording). An example of the questions is provided in Table 1.

**Table 1:**
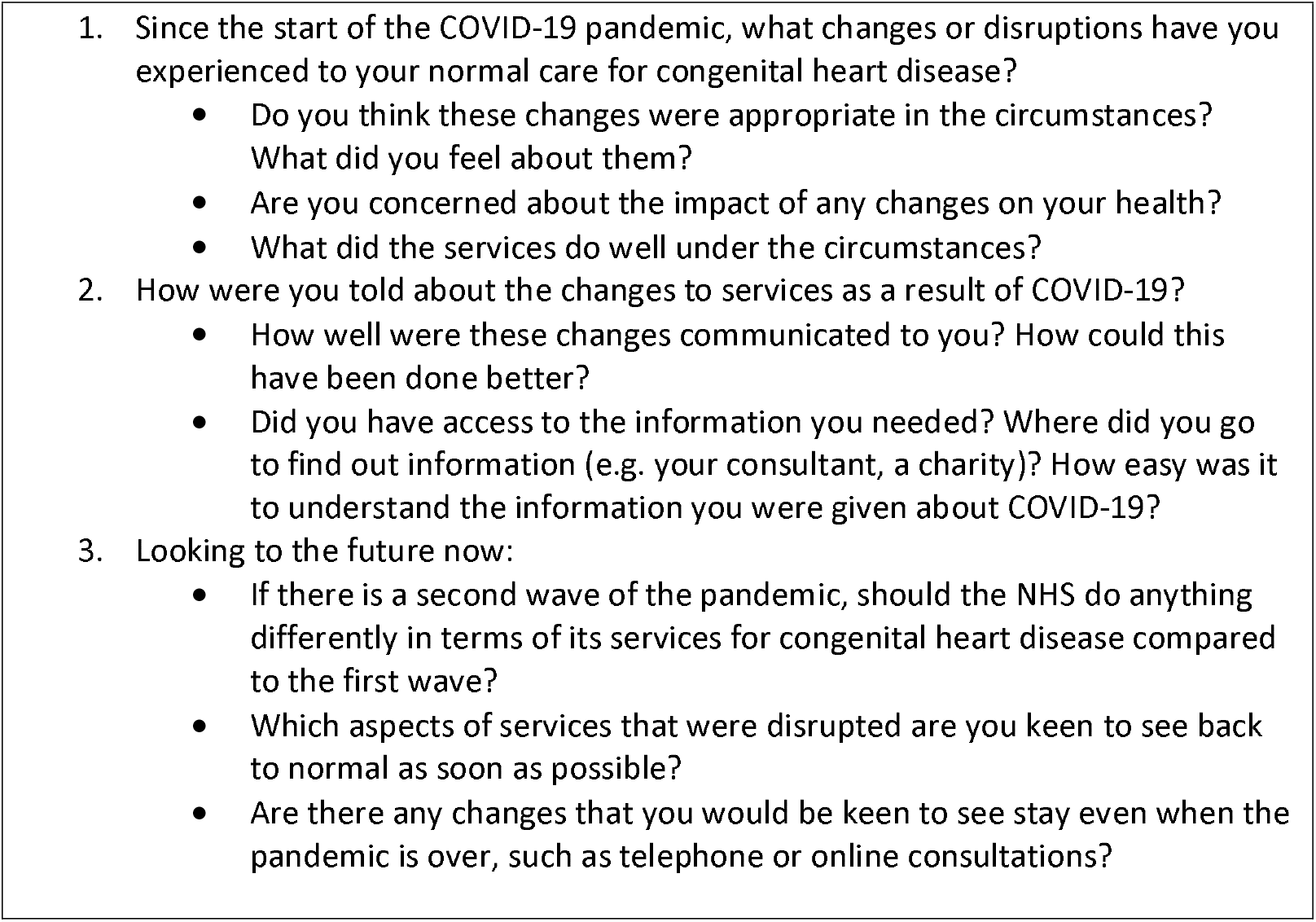
Questions for the adult patient forums. The questions for the parent/carer and teenager forums were very similar to these, with minor wording changes to reflect those respondent groups (e.g. designed to appeal to teenagers or wording appropriate for carers rather than patients).

### Data management and analysis

The charities removed any identifying details from the responses and provided the research team with a single transcript for each forum along with summary demographic details for each participant group. The transcripts were thematically analysed independently by four members of the research team (JW, SC, CP, AC). Codes were attached to segments of data, with similar codes grouped to create themes and subthemes related to the perceived impact of COVID-19 on the provision of services. The research team met to discuss the themes and subthemes and to agree the descriptive names assigned to them. The themes and suggested recommendations were then sent with the transcripts to another member of the research team (FK) to ensure that all data related to the perceived impact of COVID-19 on the delivery of services were represented appropriately in the themes.

### Ethical considerations

The Research Ethics Committee confirmed that ethical approval was not required because the forums were managed by the charities. Each charity placed privacy notices on their websites, clarifying that participants’ comments would only be visible to other members of the discussion group and the charity forum moderators and that all identifying information would be removed from discussion posts before being sent to the researchers.

## Results

Five forums were run across the three charities, with 109 participants in total. Participant demographics are shown in Table 2.

**Table 2:** Participant characteristics.

|  | Number (%) |
| --- | --- |
| Participants: Adults with CHD | 82 (75) |
| Young people with CHD | 3 (3) |
| Parents/carers of adult patients with CHD | 2 (2) |
| Parents/carers of children with CHD | 22 (20) |
| Participant gender: Male | 9 (8) |
| Female | 88 (81) |
| Unknown | 12 (11) |
| Participant age group: <16 years | 1 (1) |
| 16-20 | 2 (2) |
| 21-30 | 9 (8) |
| 31-40 | 26 (24) |
| 41-50 | 28 (26) |
| 51-60 | 24 (22) |
| >61 years | 7 (6) |
| Unknown | 12 (11) |
| Age group of person with CHD: 0-1 years | 1 (1) |
| 2-5 years | 3 (3) |
| 6-10 years | 1 (1) |
| 11-15 years | 2 (2) |
| 16-18 years | 2 (2) |
| >18 years | 82 (75) |
| Unknown | 18 (17) |
| Participant ethnicity: White | 99 (91) |
| Non-white | 0 (0) |
| Unknown | 10 (9) |
| Location of specialist service: England (North East) | 3 (3) |
| England (North West) | 8 (7) |
| England (Yorkshire and the Humber) | 3 (3) |
| England (East Midlands) | 6 (6) |
| England (West Midlands) | 16 (15) |
| England (East of England) | 3 (3) |
| England (London) | 26 (24) |
| England (South East) | 6 (6) |
| England (South West) | 9 (8) |
| Wales | 1 (1) |
| Scotland | 7 (6) |
| Northern Ireland/other | 1 (1) |
| Unknown | 20 (18) |
| Location of home: England (North East) | 3 (3) |
| England (North West) | 12 (11) |
| England (Yorkshire and the Humber) | 5 (5) |
| England (East Midlands) | 5 (5) |
| England (West Midlands) | 16 (15) |
| England (East of England) | 8 (7) |
| England (London) | 8 (7) |
| England (South East) | 13 (12) |
| England (South West) | 16 (15) |
| Wales | 4 (4) |
| Scotland | 8 (7) |
| Northern Ireland/other | 1 (1) |
| Unknown | 10 (9) |
| Complexity of CHD: Single ventricle condition | 21 (19) |
| Biventricular condition | 83 (76) |
| Unknown | 5 (5) |

\*A number of participants chose not to provide some or any demographic information

Three themes and ten subthemes related to individual condition-related factors, patient-related factors and health professional/centre factors were identified, shown in the Figure with illustrative quotes from the forums. Although there is clearly overlap between these factors, particularly in relation to communication, they represented a useful way of interpreting the data.

**Figure.**
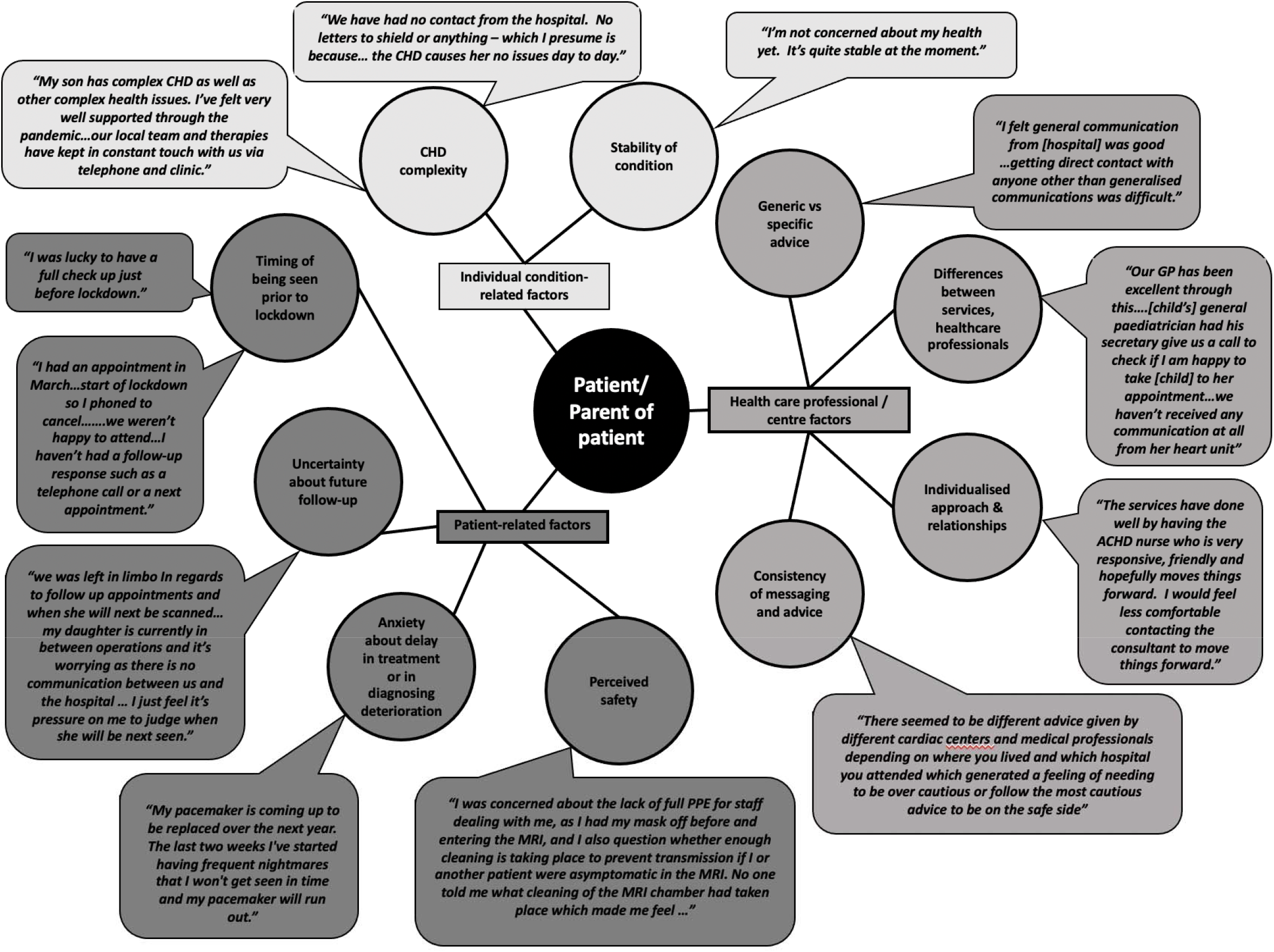
Factors influencing patients’/parents’ experiences of the impact of COVID-19 on service delivery and care

### Patient-related factors

For the majority of participants, routine clinics had been cancelled and appointments had been held via phone or video-link. Participants (both parents and patients) were largely accepting of these changes necessitated by the first wave of COVID-19 and considered them appropriate. They recognised that COVID-19 was new to everyone and that little was known about it initially, so they were mostly accepting of some of the shortcomings in communication. The timing of scheduled appointments was an important factor, with some patients seen just before the lockdown and highlighting that this was ‘*lucky*’. In contrast, others expressed uncertainty about when they would be seen and this was exacerbated if communication from their specialist centre was poor. Reported concern and/or distress were notable in patients who were newly diagnosed or who were in the process of transferring between centres: “*As I was moving from one hospital to another I had nothing [information] as neither hospital took responsibility for* me”. Some patients/parents felt that it was their responsibility to recognise signs of deterioration or the onset of problems and to decide when they or their child should be seen. Many people described the challenges of getting information about follow-up arrangements, illustrated by one patient: “*I spent many months going round in circles and being passed from pillar to post*”. Participants described feeling anxious and stressed about delays in treatment, diagnosis or identifying any deterioration in their condition. For some this stress was intensified by the loneliness brought about by the enforced isolation. A number of participants talked about safety, both in terms of perceived risks to their health from being in the hospital environment or using public transport as well as the risks of not being seen face to face and getting the necessary tests and/or interventions: *“COVID stopped me going to [hospital] for my consultation. This has its plus points and minus points. The changes under the circumstances were fine because I would have had to have travelled on public transport and it’s something that I wasn’t willing to do. However, I prefer to go to the hospital as it puts my mind at ease when they can do the necessary tests required”*.

### Individual condition-related factors

As with many other chronic health conditions, there is a spectrum of both complexity and stability of CHD and these factors seem to be important determinants of how COVID-19 was perceived to impact patients. A number of those with more complex CHD were very well supported by their specialist service as well as local primary and secondary care services. They described receiving regular phone calls and written information and, where necessary, individual arrangements for tests at local surgeries or hospitals. In contrast, some others reported having no contact from their specialist centre or guidance about whether they needed to shield and frequently felt that they had to chase for information about changes to services and guidance about shielding. However, whilst they wanted information about arrangements, those patients whose conditions were stable generally expressed low levels of concern about their health and the impact on it of any changes to their care. For patients who were unstable or who had developed new symptoms, however, the added uncertainty about how and when they might be seen was particularly stressful: “*It’s horrible knowing I have a critical illness and knowing I need surgery but not knowing how bad it is. For 5 months now I’ve been in limbo and frightened*”.

### Health care professional/centre factors

Communication was the factor that evidently had the biggest impact on patients and parents and how they perceived COVID-19 to have affected them or their child. There was general consensus that messaging and advice had been inconsistent, with different centres and different professionals offering different advice about the same thing: “*Communication from centres about shielding was very contradictory”*. Participants described variation in the contact they had had with different professionals involved in their care: some specialist centres provided excellent communication, others provided nothing; some primary and secondary health professionals were described as being exemplary *(“New GP…went over and above”)* but other patients reported having *“nothing from anyone”*. A distinction was also made between general advice and patient or condition specific advice, with the latter generally more difficult to access. Some respondents reported that clinicians, particularly cardiac specialist nurses (CLNs) who knew them/their medical history, were proactive and responsive to their queries and this was valued by patients and parents: *“I have no concerns as I find the CLNs are accessible by phone or email and I’m confident that if I had any issues I would be seen sooner”*. In contrast, others were clearly feeling very unsupported by professionals, particularly some parents who described feeling forgotten about and “*left to our own devices*.*”* What was clear, however, was the vital role played by charities in providing information and support to patients and their families, despite the acknowledged financial and other pressures that the organisations have been under.

Whilst there was a degree of acceptance and understanding about changes to services during the first wave of COVID-19, participants expressed very different expectations for managing the on-going situation and clearly articulated that, as awareness and knowledge about COVID-19 are increasing all the time, they are likely to be far less understanding and tolerant of poor communication, delays and cancellations. A number of participants expressed concerns about the big backlog of appointments and the likelihood that quite a few patients will have deteriorated, resulting in additional health issues for them and additional input and costs incurred by the NHS: *“I understand it must be very difficult but if we have a second wave I think appointments for those awaiting surgery should go ahead. I understand it’s dangerous, however leaving symptomatic patients without an appointment could be catastrophic. And would subsequently put more pressure/expense on the NHS*.*”*

## Discussion

During this study we elicited the views of a diagnostically heterogeneous group of patients with CHD and/or their parents about their experiences of changes to their specialist services as a result of COVID-19, how those changes had been communicated and what should happen in any subsequent wave of COVID-19. We identified a number of condition-related, patient-related and health professional/centre related factors that may have influenced how patients and parents/carers experienced changes to service delivery. The importance of clear, consistent communication cannot be over-estimated. A number of patients seemed to be surprised that they had not had any contact from their specialist centre, particularly those with more complex CHD, indicating that their expectations about communication with their specialist team were not met. The findings from this study suggest a somewhat mixed picture: some respondents reported being very satisfied with arrangements and described excellent communication and care; others reported some positive aspects of care delivery but they also expressed examples where communication, particularly, had been poor or inconsistent; a third group were very dissatisfied and disappointed with the lack of communication and disruption to their care. Some had a clear sense that as non-COVID-19 patients they were not a priority: *“I felt I was being ignored and that unless you were a person with COVID no-one wanted to know”*. Parents particularly expressed their concern with their experience, some of whom saw this as extending beyond cardiac-related care: *“In short, children’s care in all sectors just stopped and that is awful”*. Participants also described examples of good practice, such as the responsiveness of the clinical nurse specialists, the online support groups facilitated by psychologists and other health professionals, and the freely available YouTube educational videos developed by their consultants. One contributory factor to the different patterns of communication may have been regional levels of COVID-19 infection, with those centres in areas with high levels of infection potentially finding it harder to keep up with communication, particularly if staff were redeployed to provide front-line care in other areas.

### Limitations

Although we specifically chose a method of data collection to increase the accessibility of the research to potential participants and did achieve good diversity in terms of where participants lived and their specialist centre, participants did not reflect a broad range of ethnic groups or gender. This may be of particular salience in light of the growing body of evidence that people from black Asian and minority ethnic (BAME) groups have been disproportionately affected by COVID-19, including experiencing higher rates of mortality due to COVID-19.(12) Even if this is not shown to be the case for patients with CHD, such knowledge is likely to contribute to higher levels of anxiety in BAME individuals and may drive greater social isolation and disengagement with health care, which is an important consideration for specialist centres and the wider health service. The lack of participation from BAME groups reflects a recognised problem that they are less likely to engage with, and participate in, research than their white British counterparts (13) and speaks to the need for targeted strategies to involve, recruit and retain BAME individuals in research projects.

Charities (not limited to those who moderated the discussion forums in this research) were identified as having a vital role in providing support and information to patients and families during the first wave of COVID-19 and at times were the only perceived source of information and support. This also highlights a bigger issue of inequity as it will only be those patients and families who are willing and able (through familiarity and adequate language and literacy skills as well as internet resources) to access charity resources who will be able to benefit from them. Furthermore, many of those who are excluded from this will also be those who are less well informed and have less awareness of guidance about issues related to COVID-19. In light of the important role that they play, it may also be timely for charities to reflect on how to increase their appeal to, and membership from, BAME and other under-represented communities.

Our findings, whilst collected in relation to patients with CHD and their parents/carers, are not necessarily specific to this population and we believe reflect the experiences of many thousands of people with life-long conditions in the UK. Health care delivery changed significantly during lockdown and beyond, and as with all changes there are lessons to be learned. Drawing on what participants told us in the discussion forums, we have developed a series of recommendations (Table 3) that would, we think, improve patients’ experience of care and, potentially, their outcomes. We believe these are applicable to any patients with underlying health conditions and some, particularly those related to communication, would likely reap large benefits for relatively little input. Whilst the data were collected specifically in relation to COVID-19 and the learning has come from patients’ experiences of care during the lockdown, a number of these recommendations are relevant to the wider delivery of care to patients with chronic underlying health conditions and reflect principles of good communication and service delivery.

**Table 3:** Recommendations for improving patients’ experience of care and, potentially, their outcomes, based on what participants told us in the discussion forums. Although generated from research related to congenital heart disease, we believe that these recommendations are relevant for patients with any underlying health conditions.

|  |
| --- |
| **Communication - generic**<br>• Consistent information from all healthcare providers in relation to condition-specific advice<br>○ Includes all hospitals, GPs, community services etc<br>○ Should be routinely provided to patients with a particular condition, wherever they receive their care<br>• Produce and share information about the latest guidance and recommendations with those around the patient<br>○ Includes, but not limited to, schools, nurseries and employers<br>○ Ensure guidance is condition-specific and accessible to patients, to facilitate sharing |
| **Communication – patient specific**<br>• Clear advice and guidance about shielding (personalised to individual rather than generic)<br>○ Provided to all patients via a range of media (email, letter, easy read, text message +/- telephone)<br>• Proactive communication with patients via email or telephone<br>○ To check in with them<br>○ To update them about any changes<br>○ Determined by individual patient circumstances and need<br>• Dedicated email address/phone line with answerphone for patients to call with concerns or questions<br>○ Checked and responded to regularly by someone familiar with their individual case<br>○ Provides clear information about how frequently messages are checked and when a response can be expected |
| **Service delivery**<br>• Regular updates about services<br>○ Any curtailment of services, estimated delay times, safety precautions being put in place<br>• Greater flexibility for tests being done locally, more remote monitoring<br>• Telehealth for some/quick catch-ups or where face to face is not necessary<br>○ For communication of routine test results<br>○ Intermediate appointments for patients seen very frequently<br>○ Benefits in terms of reducing travel, time efficiency and safety<br>• Face to face where indicated/necessary<br>○ For medical tests<br>○ Where patients have complex needs<br>○ Underpinned by patient choice about how and where their care should be delivered<br>• Protection of specialist services, COVID free beds<br>• Individualised approach to patient care and follow-up<br>○ Tailored to diagnosis<br>○ Dependent on where an individual is in terms of their care pathway – e.g. waiting for a treatment intervention vs requiring routine check-up |
| **Support**<br>• Increased access to online support<br>○ Signposting to existing support groups and websites<br>• Provision of access to<br>○ Support meetings<br>○ Videos made by health professionals<br>○ Other resources established in response to COVID-19 |

## Data Availability

The nature of the qualitative data, whilst anonymised, would potentially allow for identification of participants and the data have therefore not been made available.

## Acknowledgements

The authors thank The Somerville Foundation, Little Hearts Matter and the Children’s Heart Federation for recruiting participants and moderating the forums, and for their contributions, together with the PPI group, to the development and review of the forum questions. JW is supported by the Great Ormond Street Hospital NIHR Biomedical Research Centre.

## Contributorship statement

All authors contributed to the design of the study. JW, CP, AC and SC undertook the initial analysis of the transcripts and FK independently checked that all data related to the perceived impact of COVID-19 on the delivery of services were represented appropriately in the themes. All authors contributed to the writing of the manuscript and have approved the final version.

